# Primary health center unit closures following a large-scale administrative reform: A multilevel analysis of determinants

**DOI:** 10.1101/2025.11.17.25340376

**Authors:** Visa Väisänen, Liina-Kaisa Tynkkynen, Konsta Lavaste, Timo Sinervo

## Abstract

**Background:** To address growing care demands, current health system reforms aim to strengthen primary healthcare (PHC) services and their accessibility. Simultaneously, large-scale structural reforms can lead to centralization of PHC services, yet the determinants of local service closures remain poorly understood. This study examines the determinants of PHC service point closures following a recent administrative reform in Finland, where the newly established wellbeing services counties (WSC) are currently reorganizing their local service networks.

**Methods:** Health center unit closure decisions were systematically extracted from policy documents. Publicly available municipality and WSC-level register data were utilized, encompassing population characteristics, geographics, as well as service network and reform-related factors. Multilevel logistic regression was conducted to analyze the factors associated with closures at the municipal level.

**Results:** Out of 295 municipalities, 82 were facing health center unit closures, with 45 left without a unit (previously four). A higher number of current (public) health centers (OR: 15.17, CI: 5.51–41.80), a better medical desert index value (OR: 1.90, CI: 1.27–2.83), and the WSC being a new actor with no previous joint administration (OR: 10.95, CI: 1.15–104.33) were associated with closure decisions. In contrast, greater municipal population growth (OR: 0.21, CI: 0.08–0.53) and a higher number of private clinics (OR: 0.22, CI: 0.05–0.94) were associated with lower odds of closures.

**Conclusions:** Following the administrative reform, PHC unit closures concentrated on areas with denser existing service networks, suggesting a rationalization of existing networks. Counties lacking pre-reform collaboration structures faced more extensive restructuring, emphasizing the role of administrative continuity in mitigating accumulation of service reform needs. These findings highlight how existing service configurations and prior governance structures can influence the implementation of a large-scale health system reform.

## 1. Introduction

Reforms and policy changes serve as important ways to develop and advance health systems. Reforms can be small or large in scale and slow or rapid in pace (Tuohy, 2018), and are guided by varying theoretical and strategic approaches (Oliver and Mossialos, 2005). Present health system reform needs in high-income countries are primarily driven by demographic changes, mainly ageing, which challenge both financial sustainability and the current ways to provide services (Bloom *et al*., 2015; De Biase *et al*., 2022; Rechel *et al*., 2009). Additionally, the growing burden of chronic diseases (Hacker, 2024), external and internal crises (Berardi *et al*., 2024), and advancements in medicine (Bodenheimer, 2005) pose challenges to health systems’ resilience. In European countries with universal health systems, the aims of reform planning and implementation have focused on improving the efficiency and, more recently, the quality of care (Hernández-Quevedo *et al*., 2018). Reforms have especially concentrated on strengthening primary healthcare (PHC), changing governance structures, and centralizing hospital networks (Polin *et al*., 2021). Previous discussions on re-centralization and decentralization of healthcare services (Saltman, 2007) are accelerating, with countries attempting to balance local know-how and national governance aims emphasizing efficiency, equity, and sustainability (e.g., (Burke *et al*., 2018; Dubas-Jakóbczyk *et al*., 2023; Sundhedsstruktur komissionen, 2024)).

Health system reforms can have implicit or explicit repercussions for PHC service networks. For instance, health system reforms after the 2008 financial crisis focused on re-centralization and cost containment (Berardi *et al*., 2024), which had widespread negative effects on availability of care, mainly due to longer waiting times and higher out-of-pocket payments, but also due to changes in service networks (Doetsch *et al*., 2023; Stuckler *et al*., 2017). In England, encouraged financially by the National Health Service long term plan (Alderwick and Dixon, 2019; Parkinson *et al*., 2021), general practitioner (GP) practices have formed into larger primary care networks (Morciano *et al*., 2020) and simultaneously many practices are continuing to face closures (Hutchinson *et al*., 2023). These reforms highlight the pursuit of economies of scale by utilizing larger care units and multidisciplinary practices. The trend towards more centralized PHC provision is further accelerated by the growing private equity investment in the European health sector, which has the potential to increase market consolidation (Rechel *et al*., 2023). PHC service networks can also be affected by more incremental reforms, such as development of provider incentives (Flinterman *et al*., 2023; Holte *et al*., 2015) and quota-based systems (Ozegowski, 2013). For example, while the Swedish patient choice reform initially concentrated PHC services towards more affluent urban areas, certain provider incentives have been successful in attracting clinics also to underserved rural areas (Fredriksson and Isaksson, 2022; Janlöv *et al*., 2023). In health systems with strong reliance on public provision of PHC, service networks can be steered and reformed through direct centralized planning and decision-making. However, this task concurrently grapples with the realities of cost-containment and efficiency aims, politics, and workforce availability.

The current reform trends, in tandem with the continuing urbanization (“Urbanization in Europe 2000–2018”, 2024) and peri-urbanization (Shaw *et al*., 2020), are likely resulting in an increasing frequency of PHC center or GP clinic closures or mergers across Europe in the coming years. Recent literature examining practice closures in PHC has largely focused on outcomes such as service utilization and population health impacts. Research has found the remaining GP practices growing in size (Hutchinson *et al*., 2023), a decline in patient satisfaction (Hutchinson *et al*., 2023), and a potential shift towards emergency and acute services, especially among vulnerable groups (Palacios *et al*., 2024). Literature also suggests that most clients successfully transition to new services with no perceived differences in quality of care (Trombetta *et al*., 2025), and that the newly re-matched clients might begin treatments due to a reassessment of care needs, potentially leading to improved care and health outcomes (Simonsen *et al*., 2021).

Despite the growing trend of reforming PHC service networks, the determinants of PHC unit closures have been scarcely examined, and thus, the factors guiding and influencing local-level service planning remain unidentified. This is of special relevance in tax-funded health systems, where public stewardship and local policymakers can directly decide the service networks. The recent administrative health system reform in Finland and the subsequent extensive local service network reconfiguration enable examining the decision-making processes and determinants of PHC center unit closures.

The Finnish health and social care reform came into effect in the beginning of 2023. Previously PHC and social services were organized by the 309 municipalities (or joint authorities formed by the municipalities) and specialized care by 20 hospital districts, both funded mainly by municipal taxes and government subsidies. The 2023 reform transferred all the previously fragmented responsibilities to 21 new autonomous wellbeing services counties (WSC) governed by elected council members and funded almost entirely by the national government. The capital city of Helsinki (hereafter, referred to as a WSC) continues to organize its own services. In the capital region (Uusimaa), the specialized care for the five WSCs in the area is organized jointly by the HUS Group joint authority. The reform goals were to ensure equal services for all residents, improve the accessibility of services, and to curb the growth of the costs. However, the reform implementation has been challenging, with virtually all WSCs facing large budgetary deficits caused by multiple factors, including inflation, IT and pay harmonization, and a historic underreporting of costs by municipalities (Tynkkynen *et al*., 2024). As the funding comes from the government, budget deficits must be covered through expenditure cuts, resulting in widespread saving plans. Simultaneously, the WSCs inherited the PHC service networks designed for the system run by the municipalities. Therefore, one of the first tasks of the WSCs was to adapt their service networks to accommodate the new administrative structure, which includes centralization and closures of health center units.

Health centers are the main providers of comprehensive primary care services in Finland. They consist of larger centers and smaller units, in which salaried GPs and nurses provide a varying set of primary care services that all residents in Finland are entitled to (Keskimäki *et al*., 2019). The services provided differ significantly between larger healthcare and social welfare centers and smaller units, of which the latter might offer only limited ambulatory services. Pre-reform service provision centered around health center units, with virtually all municipalities having at least one. In the Finnish PHC, nurses play a major role in providing care, and they often act as gatekeepers for seeing a physician, who then act as gatekeepers to specialist care. Due to the occupational healthcare scheme and voluntary private health insurance, the population served by the health centers is biased towards those outside employment, pensioners, and people with lower socioeconomic status. Despite this, the centers remain a central pillar of PHC services in Finland, which in addition to outpatient medical services, provide multidisciplinary care for chronic diseases and mental health and substance use (Keskimäki *et al*., 2019).

The planned closures have sparked lively discussion among policymakers and the public. As health center units are both historically and culturally rooted in the municipalities’ identities, many residents fear that planned closures signal the withering of services in general (Kihlström *et al*., 2025). The WSCs have justified and emphasized the necessity of reorganizing the PHC service networks with aims of modernizing services and ensuring the sufficiency of health workforce. Meanwhile, other WSCs have opted to maintain their existing service networks. The situation presents a unique opportunity to investigate decision-making following a large-scale administrative reform and gather evidence of the determinants of PHC center closures. In response to the gap identified in the research literature, we aimed to exploit the ongoing reform implementation to analyze policy and decision-making related to health center unit closures. Two research questions were formulated:

1. Which municipality- and WSC-level factors are associated with the decision to close a health center unit in the area of a municipality?
2. Which factors explain the variation in closure decisions between WSCs?

## 2. Materials and methods

### 2.1 Study design

This is an observational study utilizing a multilevel modeling strategy.

### 2.2 Data collection

Publicly available aggregated register data were retrieved from Statistics Finland, National Land Survey of Finland, and the Finnish Institute for Health and Welfare. WSC websites were accessed for various policy documents, including strategies, service network analyses, and official meeting records and decisions.

The resulting cross-sectional dataset included all Finnish municipalities (excluding those in the autonomous Åland islands) in 2025 (n = 292) as observation units. Additionally, to accommodate for the multilevel structure, the capital city of Helsinki (treated as a WSC) was divided into eight official subdivisions (treated as municipalities) with their own data points.

### 2.3 Outcome variable

The outcome measure was the planned closure of a health center unit in the area of a municipality, with 1 indicating that at least one health center unit has been or will be closed, and 0 meaning that no closures are planned. Closures were determined by systematically reviewing the WSCs’ websites, strategies, service network analyses, as well as WSC councils’ meeting notes and decisions, with the latest check conducted on June 10, 2025.

In many cases, the WSCs reduced service networks by downsizing the service portfolios of the health center units rather than closing them entirely. We chose to determine closure based on ambulatory outpatient care: the unit was not identified as closed if a physician was planned to be present at least once a week and a nurse multiple times a week. Additionally, we took the uncertainty of decision making into account by marking closures lacking a political decision or planned for later than 2027 (five years after the reform) as unclear and examining them in the sensitivity analysis.

### 2.4 Explanatory variables

In accordance with the research literature and the reform implementation, we identified several potential municipality and WSC-level elements which could influence decision-making, presented in our conceptual framework (Figure 1). As the decision-making occurs in the WSCs, their characteristics and conditions (e.g., fiscal situation) are central. Although municipalities no longer play a role in the health system, the pre-reform service networks were inherited from them, and consequently their role and population and geographical characteristics are still of interest. These municipality and WSC-level factors were supplemented by the overall context, including pre- and post-reform conditions, in addition to previous PHC closures.

**Figure 1:**
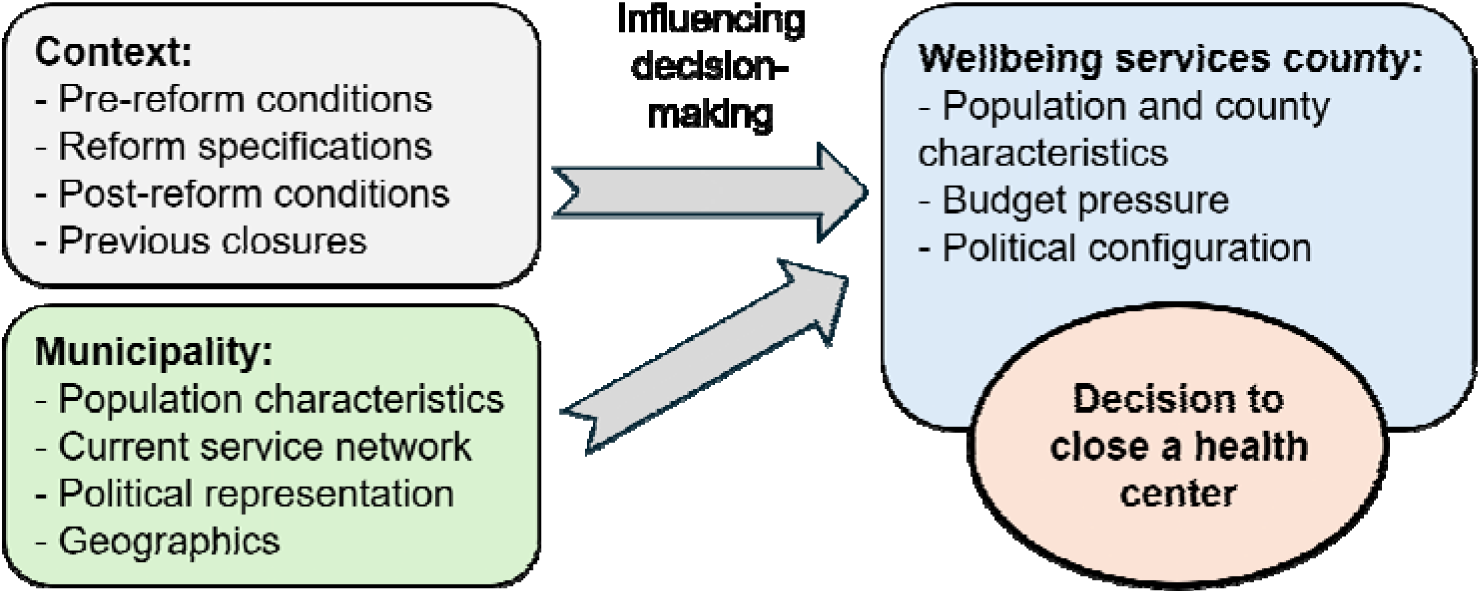
Conceptual framework of the study illustrating the factors potentially influencing health center unit closures.

The municipality-level variables consisted of population and geographical characteristics, current service network, and political representation. Population in year 2023, population density (1000 per km²), population growth (%) from 2013 to 2023, and the proportion of residents aged 65 or older were included for population characteristics. Geographical characteristics (in addition to population density) included the previously constructed Finnish medical desert index (Väisänen, Satokangas, *et al*., 2025), which comprises needs adjusted care supply data and average travel times to the nearest health center unit. The current service network was characterized by the number of health center units (mostly WSC-owned, but some provision purchased from private providers) and private clinics (purely private services), as well as the number of health center unit closures in the last 10 years. Lastly, to measure for the municipalities’ political representation in the regional decision-making, the percentage of WSC councilmembers from the municipality was used. Weaker spatial political representation has previously been linked to the closure of public services (Harjunen *et al*., 2023).

The WSC-level variables consisted of population and WSC characteristics, economic situation, and political configuration. Additionally, pre- and post-reform conditions, including reform specifications, were included. WSC characteristics included population density (1000 per km²) and the number of municipalities within the WSC. Some municipalities had organized PHC services jointly pre-reform and might consequently have already optimized their networks and retained collaboration structures. Therefore, a patchwork county indicator was included (1 = most municipalities in the WSC previously organized their care jointly, 0 = no previous cooperative service provision). Additionally, the presence of a university hospital (1 = yes) and the capital region (1 = yes), where specialized care is organized jointly (by the HUS Group) rather than WSC-wise, were included as cooperative specialized care provision may influence decision-making regarding the PHC service networks. The overall budgetary deficit or surplus at the end of year 2024 (€ per person) was used to measure the level of austerity, which could drive closure decisions (Doetsch *et al*., 2023; Stuckler *et al*., 2017). Lastly, to measure the political configuration and power, the seat share of the largest political party was included, with a hypothesis that stronger political blocks could be linked to better decisiveness and governability of decision-making.

#### Statistical analysis

Descriptive statistics were used to compare municipalities by health center unit closure status. To examine the determinants of closures, multilevel logistic regression was performed, with a municipality facing health center unit closure or closures as the dependent binary variable. The municipalities (first level) were nested within the WSCs (second level). All continuous independent variables were standardized (Z-score). There was no missing data. Satterthwaite’s degrees of freedom method was used to calculate p-values (Satterthwaite, 1941) and Akaike information criterion were used to assess model performance. Data management and statistical analyses were performed using R Statistical Software version 4.2.3 for Windows 11 (R Core Team, 2020) and packages lme4 (Bates *et al*., 2003) and lmerTest (Kuznetsova *et al*., 2013). Sensitivity analysis was conducted by including the unconfirmed closure of health center units in addition to a linear modeling strategy (see Supplementary file).

To determine how different factors explain the variation in the local and the WSC-level, a stepwise approach was utilized. First, a null model (0) was constructed. Next, municipality-level variables measuring population characteristics (1) and current service network (2) were added sequentially. In models 3-4, WSC-level reform conditions (3) and the characteristics (4) were incorporated. In the final model, political factors (5) were included.

## 3. Results

The population, geographics, service network, and reform-related characteristics of the municipalities and WSCs are presented in the Supplementary file. Both the municipalities and WSCs had significant variation in population characteristics and the current PHC service networks. The number of health center units in the area of municipalities ranged from 1 to 12, with a mean of 1.7. Over half of the WSCs were patchwork counties (no pre-reform collaboration in PHC), while one fifth had a university hospital or were part of the HUS Group (the capital region).

The current Finnish PHC service network and the planned closures are illustrated in Figure 2. The closures appear to concentrate in western Finland, particularly in patchwork counties. In the areas with the lowest population densities, especially northern Finland, very few closures were planned.

**Figure 2:**
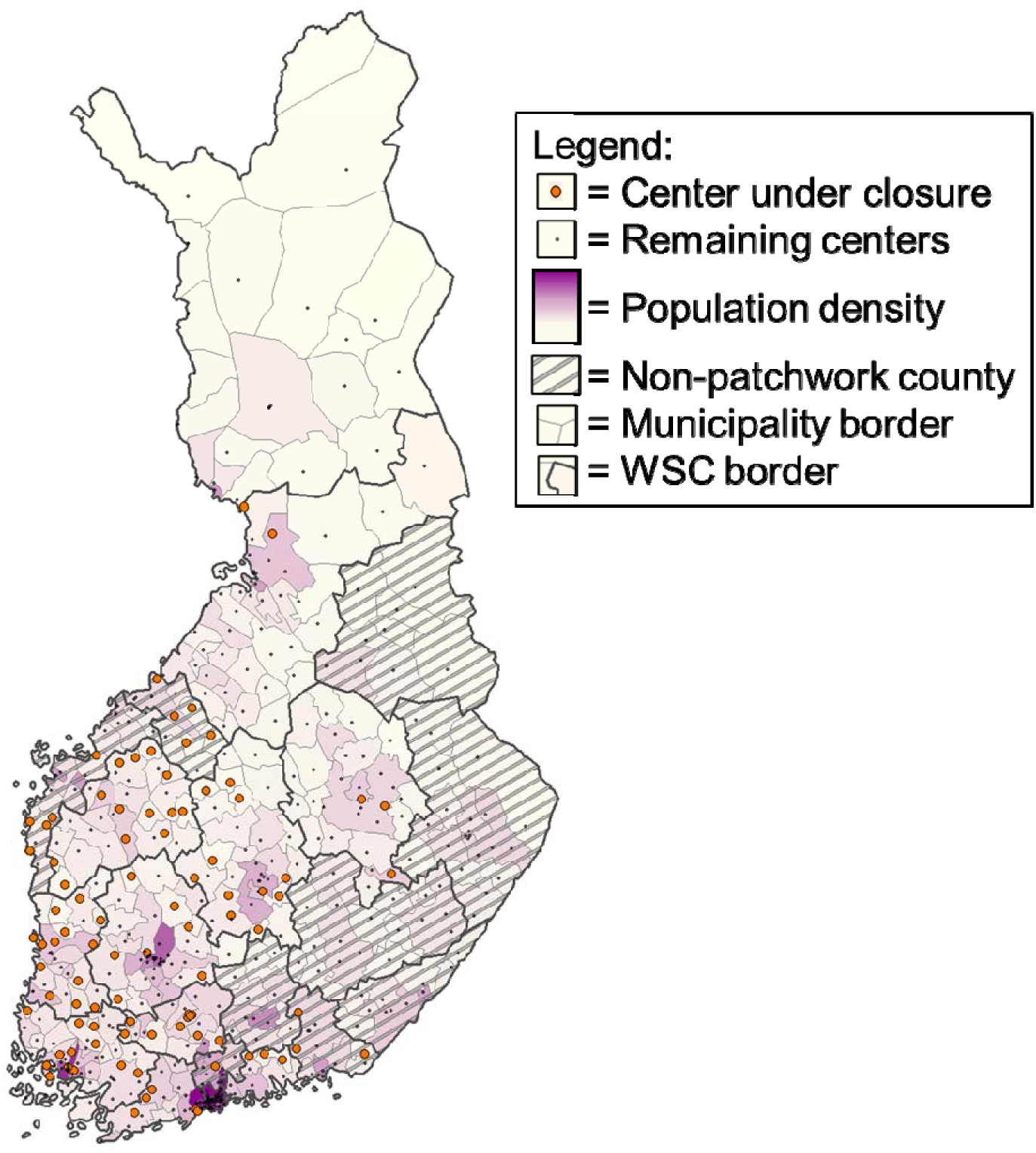
Map of Finnish primary health center unit closures (n = 108 in 82 municipalities). Patchwork county refers to wellbeing services counties (WSC) without pre-reform collaboration (such as joint administration of municipalities). Private clinics are not shown.

Next, municipalities with and without planned PHC center unit closures were compared (Table 1). Municipalities with planned closures had on average higher population sizes (28,562 vs. 15,049) but lower population densities (54.6 vs. 178.0). Their current service networks were denser, with better medical desert index values (0.36 vs. −0.12) and more public and private clinics (2.6 vs. 1.3 and 1.5 vs. 0.9, respectively). A significant proportion of closures were in municipalities located in patchwork counties (86.6%). The number of municipalities without a health center unit was 4 prior to the reform, rising to 45 after the confirmed PHC center unit closures and to 61 when including unconfirmed closures.

**Table 1:** Comparison of characteristics between municipalities with and without planned health center unit closures in their area (n = 295).

|  | Municipalities with planned PHC center unit closures (n = 82) |  |  | Municipalities with no PHC center unit closures* (n = 213) |  |  |
| --- | --- | --- | --- | --- | --- | --- |
|  | Mean / % | SD | Range | Mean / % | SD | Range |
| **Population characteristics** |  |  |  |  |  |  |
| Population | 28,562 | 57,129 | 682–314,024 | 15,049 | 26,908 | 912–247,443 |
| Population density (per km<sup>2</sup>) | 54.6 | 151.8 | 1.4–1,005.3 | 178.0 | 786.6 | 0.2–6,827.0 |
| Population aged 65 or more (%) | 29.5 | 6.7 | 15.1–43.6 | 31.0 | 7.9 | 11.4–47.2 |
| Population growth 2013–2023 (%) | -8.5 | 10.8 | -27.2–17.0 | -8.3 | 10.1 | -29.7–16.7 |
| **Service network** |  |  |  |  |  |  |
| Medical desert index, physical | 0.36 | 1.21 | -1.96–5.99 | -0.12 | 0.86 | -1.96–2.30 |
| Number of public health center units | 2.6 | 2.5 | 1–12 | 1.3 | 1.0 | 1–8 |
| Number of private clinics | 1.5 | 3.1 | 0–18 | 0.9 | 1.8 | 0–16 |
| **WSC, reform and administration** |  |  |  |  |  |  |
| Councilmembers from the municipality (%) | 9.6 | 16.1 | 0–69.5 | 6.6 | 11.0 | 0–87.0 |
| Previous closures of health center units in municipality | 0.22 | 0.65 | 0–4 | 0.19 | 0.53 | 0–3 |
| In patchwork counties (%) | 86.6 |  |  | 62.9 |  |  |
| County has a university hospital (%) | 32.9 |  |  | 36.6 |  |  |
| In HUS Group area (%) | 9.8 |  |  | 11.3 |  |  |
| * Excluding the municipalities of Kaustinen, Korsnäs, Nousiainen, and the Östersundom major district of Helsinki, which had no health center units. |  |  |  |  |  |  |

Results of multilevel modeling, conducted in six steps, are presented in Table 2. A large proportion of the variance in PHC center unit closures in the areas of municipalities was explained by differences at the WSC level (ICC: 0.36). Models 1 and 2, adding municipality characteristics and service network factors, did not significantly change the variance explained by the group level. Model 3 (ICC: 0.27), which included WSC-level reform conditions, however, explained nearly a third of the variation in the regional level, with the WSC being a patchwork county especially emerging as a predictive factor. Policy and political factors (model 5) had marginal effect on the proportion of variation explained by the fixed effects. Strong unexplained group-level variance remained in the final model (ICC: 0.25).

**Table 2:** Results of the multilevel logistic regression analysis, with municipalities (level 1) clustered within the wellbeing services counties (level 2). The outcome variable was the decision to close a health center unit (minimum one) in the area of a municipality (0/1).

|  | Null model | Model 1: Municipality characteristics | Model 2: Municipality service network | Model 3: WSC reform conditions | Model 4: WSC characteristics | Model 5: Policy and political factors |
| --- | --- | --- | --- | --- | --- | --- |
|  |  | OR (95% CI) | OR (95% CI) | OR (95% CI) | OR (95% CI) | OR (95% CI) |
| **Population** |  | **4.98 (2.07–12.00)\*\*\*** | 2.59 (0.45–14.99) | 2.20 (0.37–13.05) | 1.79 (0.29–10.96) | 2.81 (0.28–27.85) |
| **Population density (per km<sup>2</sup>)** |  | **0.01 (0.00–0.49)\*** | 0.11 (0.01–1.18) | 0.15 (0.01–2.14) | 1.28 (0.04–41.49) | 0.90 (0.01–74.74) |
| **Proportion of population aged 65 or older (%)** |  | **0.48 (0.23–0.99)\*** | **0.39 (0.17–0.92)\*** | **0.41 (0.18–0.95)\*** | **0.41 (0.18–0.95)\*** | **0.41 (0.17–0.96)\*** |
| **Population growth (2013–2023)** |  | **0.33 (0.15–0.71)\*\*** | **0.19 (0.08–0.48)\*\*\*** | **0.20 (0.08–0.50)\*\*\*** | **0.20 (0.08–0.49)\*\*\*** | **0.21 (0.08–0.53)\*\*** |
| **Number of public health center units** |  |  | **12.50 (4.95–31.59)\*\*\*** | **12.94 (5.16–32.47)\*\*\*** | **14.26 (5.45–37.29)\*\*\*** | **15.17 (5.51–41.80)\*\*\*** |
| **Number of private clinics** |  |  | **0.17 (0.04–0.70)\*** | **0.18 (0.04–0.71)\*** | **0.17 (0.04–0.68)\*** | **0.22 (0.05–0.94)\*** |
| **Medical desert index** |  |  | **1.97 (1.32–2.92)\*\*\*** | **1.95 (0.31–2.90)\*\*\*** | **1.92 (1.29–2.85)\*\*** | **1.90 (1.27–2.83)\*\*** |
| **Previous health center unit closures in the municipality (10 years) (1)** |  |  | 0.67 (0.19–2.38) | 0.73 (0.20–2.63) | 0.72 (0.20–2.63) | 0.69 (0.18–2.69) |
| **WSC: Patchwork county (1)** |  |  |  | **5.62 (1.15–27.43)\*** | **10.60 (1.41–79.81)\*** | **10.95 (1.15–104.33)\*** |
| **WSC: University hospital (1)** |  |  |  | 0.49 (0.09–2.68) | 1.02 (0.11–9.15) | 0.60 (0.05–7.59) |
| **WSC: HUS-area (1)** |  |  |  | 1.72 (0.22–13.37) | 1.39 (0.10–19.46) | 1.05 (0.07–15.69) |
| **WSC: Population density** |  |  |  |  | 0.08 (0.00–7.45) | 0.05 (0.00–17.53) |
| **WSC: Number of municipalities** |  |  |  |  | 0.50 (0.13–1.97) | 0.48 (0.11–2.04) |
| **WSC: Budget surplus or deficit per person** |  |  |  |  |  | 1.36 (0.59–3.13) |
| **Councilmembers from the municipality (%)** |  |  |  |  |  | 0.53 (0.19–1.44) |
| **Largest party seat share (%)** |  |  |  |  |  | 0.91 (0.44–1.88) |
| Variance of the random intercept | 1.86 | 2.05 | 1.94 | 1.24 | 1.14 | 1.09 |
| ICC | 0.36 | 0.38 | 0.37 | 0.27 | 0.26 | 0.25 |
| Δ ICC |  | 0.02 | -0.01 | -0.10 | -0.01 | -0.01 |
| AIC | 322.4 | 297.5 | 249.5 | 250.2 | 251.6 | 255.1 |
| *** = p < 0.001, ** = p < 0.01, * = p < 0.05<br>OR = Odds ratio, WSC = Wellbeing services county, ICC = Intraclass correlation coefficient, AIC = Akaike information criterion<br>Note: Municipalities Kaustinen, Korsnäs, Nousiainen, and the Östersundom major district of Helsinki had no health centers and were thus excluded from the analyses. |  |  |  |  |  |  |

At the municipality-level, the number of health center units (OR: 15.17, CI: 5.51–41.80) and a better medical desert index value (OR: 1.90, CI: 1.27–2.83) were positively associated with closure decisions. Higher population growth (OR: 0.21, CI: 0.08–0.53), the number of private clinics (OR 0.22, CI: 0.05–0.94), and a higher proportion of population aged 65 or older (OR: 0.41, CI: 0.17–0.96) were associated with lower odds of PHC center unit closure decisions. On the WSC-level, the municipality being in a patchwork county (OR: 10.95, CI: 1.15–104.33) was a strong predictor of closures.

WSC characteristics and political factors, such as the number of municipalities, budget deficit, or councilmembers being from the municipality, were not associated with closures. We also examined the effects of different political parties but found no evidence of association with closures. Inclusion of political party seat proportions significantly reduced model performance and were therefore omitted.

## 4. Discussion

We investigated the planned PHC center unit closures following the administrative health system reform in Finland. The closures focused on areas with declining populations but denser existing service networks, indicating a general rationalization of service networks. Regardless, 45 municipalities (out of 295) will be left without a health center unit (previously four). As the closures mostly concentrated on urban or peri-urban areas, the previously identified medical deserts appear to have been considered. As patchwork counties had significantly higher odds of closures, the role of administrative continuity in accumulated reform needs is emphasized. In the final model, significant variation in closure decisions remained between the WSCs, indicating the presence of unexplored determinants such as the workforce situation, which could further explain the differences.

Our study presents evidence of three main determinants driving PHC center unit closure decisions, highlighting the general rationalization of service networks. Municipal demographics, particularly population growth and age structure, were associated with closure decisions. This suggests that anticipated future care needs has been a factor in decision-making, which is important in the continuously urbanizing and ageing Europe (Bloom *et al*., 2015; “Urbanization in Europe 2000–2018”, 2024). Second, the current service networks appear central in decision-making, with both the number of health center units and private clinics, in addition to medical desertification, being associated with closure decisions. While this result was expected, the fact that underserved rural areas were considered is a positive indication that the closures may not exacerbate existing inequalities in care access. Third, the WSC being a patchwork county was strongly associated with increased closure decisions, suggesting that counties with administrative continuity have likely already optimized their service networks, resulting in both a lower need to restructure their service networks and a fewer number of units to potentially close. This result highlights the importance of administrative continuity in mitigating accumulated service reform needs.

The political factors analyzed were not associated with closure decisions, contrary to expectations for a politically driven process. Previous qualitative assessments of the Finnish reform implementation have included stakeholders voicing concerns about the limited (political) decision-making capability and rigid, yet disjointed and fragmented, central steering by multiple ministries (Paananen *et al*., 2024; Tynkkynen *et al*., 2025). This is in line with our findings, as the general rationalization of service networks appears primarily driven by geographical, population, and service network factors, with little to no room for political decision-making. Policy innovation and diffusion, which promote both the quality and efficiency of care, are often seen as the benefits of local-level decision-making (Costa-Font and Ferrer-i-Carbonell, 2022; Costa-Font and Turati, 2018). These positive effects might not emerge if diverse policy innovation is not encouraged, for instance due to the inability of the WSCs to collect extra revenue. While our findings might be explained by the extensive and sudden restructuring process induced by the challenging operating environment (Tynkkynen *et al*., 2024), the credibility of the new self-governing WSCs might affected if the preferences of the populations and decision-makers are not fully reflected in the policy decisions.

Internationally, our findings are unique, as no comparable administrative health system reforms have been implemented in recent history, and evidence on previous PHC center unit closures remains scarce. As many countries are contemplating health system reforms, including centralization and consolidation of actors and service networks (Hernández-Quevedo *et al*., 2018; Polin *et al*., 2021), our results have implications for future reform planning and implementation. For instance, both Ireland and Denmark are introducing new regional actors (Burke *et al*., 2018; Sundhedsstruktur komissionen, 2024). Structural reforms can facilitate the reconfiguration of PHC services, which can promote policy innovation (Islam, 2021), potentially leading to a rationalization of service networks and more equitable service provision. If the reforms result in service consolidation, the existing service networks and underserved areas should be considered to ensure that closures are targeted as appropriately as possible. Health systems relying on private GP practices may need to integrate provider incentives (Ozegowski, 2013) to guarantee adequate service networks in rural or underserved areas. Reforms which include transferring responsibilities of service provision, promoting administrative continuity, for instance with phased transitions or other pre-reform collaborative structures, can mitigate the accumulation of service needs and the need for extensive and sudden service network configuration.

Following the closure of PHC center units, it remains essential to monitor the newly configured service networks, including costs, quality, accessibility, and availability of care. This includes analysis of services adjacent to PHC (e.g., long-term care), which have also been undergoing changes in their service networks. While the planned closures appear to mainly target areas with denser service networks, currently underserved communities still continue to suffer from poor care access (Väisänen, Satokangas, *et al*., 2025). Access to rural PHC services could be improved for instance by increased use of mobile clinics, telehealth services, and greater integration of non-profit service provision (Gizaw *et al*., 2022). The WSCs are accelerating efforts to implement and strengthen digital services, with 49 % of all public PHC visits being done remotely for nurses and 17 % for physicians (excluding consultations between professionals) in 2024 (Finnish Institute for Health and Welfare, 2025). However, poor digital competence, common especially among older people, can hinder care utilization and lead to digital exclusion (Heponiemi *et al*., 2022, 2024), suggesting that substitutive digital care pathways may not be appropriate or sufficient for all populations. Additionally, the use of mobile clinics and service points with limited services has increased. Many of the units were not fully closed, but the range of services offered was significantly reduced (e.g., no walk-in services). While this may help maintain some level of service provision, especially for chronic care clients, it remains critical to ensure that the remaining health center units can adequately meet the needs of local populations.

## Limitations

The study focuses on the early phases of the administrative reform and the initial changes made thereafter, and therefore, long-term effects of the reform are beyond the scope of our investigation. Next, as the PHC provision in Finland is based on administratively determined health center units, the generalizability of our findings to systems based on independently established GP practices might be limited. Furthermore, although various dimensions were included in our modelling strategy, it is likely that additional factors explain the variation between WSCs, which remained high (ICC: 0.25 in the full model). For instance, we could not include information on the workforce availability, as no comprehensive data were available. Moreover, the administrative reform of Finland is unique in its breadth, and consequently more incremental reforms may involve different determinants in service network decisions. Nonetheless, the present study provides novel insights into PHC center closures and the related decision-making following a large-scale administrative health system reform.

## Conclusions

Large-scale administrative health system reforms can result in significant reconfiguration of PHC service networks, including closures of local service points. Our findings indicate that the planned closures in Finland focused on areas with denser existing service networks and declining populations, indicating a general rationalization of service networks. Additionally, areas facing poor accessibility and availability of services were considered. Patchwork counties (i.e., counties without pre-reform collaboration structures) planned a greater number of closures, suggesting that administrative discontinuity is linked to greater accumulated service reform needs. These findings emphasize the importance of existing service configurations and pre-reform governance structures in determining how large-scale reforms are implemented at the local level. Future administrative health system reforms should account for existing service networks and promote administrative continuity to reduce the need for extensive and sudden post-reform local service network changes.

## Data Availability

The datasets used and/or analyzed during the current study are available from the corresponding author on reasonable request.

## List of abbreviations

GP: General practitioner
PHC: Primary healthcare
WSC: Wellbeing services county

## Acknowledgements

None.

## Declarations

### Funding

This research was co-funded by the Research Council of Finland (grant number 354745).

### Conflict of Interest statement

The authors report there are no competing interests to declare.

### Authors’ contributions

VV: Conceptualization, writing – original draft, methodology, formal analysis. L-KT: Writing – review and editing. KL: Writing – review and editing. TS: Writing – review and editing, supervision. All authors read and accepted the final manuscript.

### Ethics approval and consent to participate

The study utilized publicly available routinely collected aggregated, and thus anonymous, municipality and WSC level data, and as such, ethical approval by an ethics committee was not needed.

### Consent for publication

Not applicable.

### Prior dissemination

The manuscript draft has been posted to the medRxiv preprint server (Väisänen, Tynkkynen, *et al*., 2025). A digital poster containing preliminary findings from this study was available to attendees at the 18^th^ European Public Health Conference in 2025 (Väisänen and Sinervo, 2025).

## Notes

### Competing Interest Statement

The authors have declared no competing interest.

### Summary of Updates

Abstract, introduction, and discussion updated.

