## Supplementary file for "Primary health center unit closures following a large-scale administrative reform: A multilevel analysis of determinants"

**Annex 1: Municipality and wellbeing services county characteristics.**

|  | **Municipalities (n = 295)** | | | **Wellbeing services counties** **(n = 22)** | | |
| --- | --- | --- | --- | --- | --- | --- |
|  | **Mean/%** | **SD** | **Range** | **Mean/%** | **SD** | **Range** |
| **Population characteristics** |  |  |  |  |  |  |
| Population | 18,596 | 37,970 | 682–314,024 | 253,332 | 165,903 | 67,736–674,500 |
| Population density (per km^2^) | 142.2 | 670.6 | 0.2–6,827.0 | 220.0 | 689.7 | 1.9–3,144.4 |
| Population aged 65 or older (%) * | 30.5 | 7.7 | 11.4–47.2 | 25.2 | 4.5 | 16.3–33.4 |
| Population growth 2013–2023 (%) * | -8.4 | 11.4 | -29.7–17.0 | -0.1 | 6.9 | -10.6–14.9 |
| **Service network** |  |  |  |  |  |  |
| Medical desert index, physical * | 0.02 | 1.00 | -1.96–5.99 | 0.04 | 0.54 | -1.09–0.95 |
| Number of public health center units | 1.7 | 1.7 | 1–12 | 22.7 | 12.8 | 8–54 |
| Number of private clinics | 1.1 | 2.2 | 0–18 | 14.6 | 9.2 | 4–39 |
| **WSC, reform and administration** |  |  |  |  |  |  |
| Number of municipalities |  |  |  | 13.6 | 7.4 | 2–30 |
| Councilmembers from the municipality (%) | 7.4 | 12.6 | 0–87.0 |  |  |  |
| Patchwork counties (%) | 69.5 |  |  | 54.5 |  |  |
| University hospital (%) | 35.6 |  |  | 22.7 |  |  |
| HUS Group (the capital region) (%) | 10.8 |  |  | 22.7 |  |  |
| Previous closures of health center units | 0.19 | 0.56 | 0–4 |  |  |  |
| WSC budget deficit per person (€) |  |  |  | -185.0 | 122.0 | -422.7–5.4 |
| WSC largest party seat share (%) |  |  |  | 32.2 | 7.3 | 22.8–54.2 |
| * Wellbeing services county information calculated using population-weighed aggregates from the municipality-level data.  Note: The major districts of Helsinki were included in the municipalities, and the city of Helsinki was treated as a wellbeing services county. Municipalities of Kaustinen, Korsnäs, and Nousiainen, as well as the Östersundom major district of Helsinki had no health center units and were consequently excluded. | | | | | | |

Sensitivity analysis was conducted by including the unconfirmed closure of health center units (Annex 2) in addition to a multi-level linear modeling strategy (Annex 3).

**Annex 2: Full models of confirmed closures only (1) and including closures which are either unconfirmed, still under decision-making, or planned beyond year 2027 (2).**

| **Full model comparison:** | **1. Confirmed closures only (n = 82)** | **2. Including unconfirmed closures (n = 109)** |
| --- | --- | --- |
|  | **OR (95 % CI)** | **OR (95 % CI)** |
| **Population** | 2.81 (0.28 – 27.85) | 1.29 (0.21 – 7.88) |
| **Population density (per km2)** | 0.90 (0.01 – 74.74) | 1.65 (0.69 – 3.94) |
| **Proportion of population aged 65 or older** | **0.41 (0.17 – 0.96)*** | 0.55 (0.27 – 1.14) |
| **Population growth (2013-2023)** | **0.21 (0.08 – 0.53)**** | **0.30 (0.14 – 0.68)**** |
| **Number of public health centers** | **15.17 (5.51 – 41.80)***** | **23.70 (7.73 – 72.65)***** |
| **Number of private clinics** | **0.22 (0.05 – 0.94)*** | 0.42 (0.16 – 1.07) |
| **Medical desert index** | **1.90 (1.27 – 2.83)**** | **1.50 (1.05 – 2.15)*** |
| **Previous closures in municipality (10 years) (1)** | 0.69 (0.18 – 2.69) | 0.84 (0.22 – 3.16) |
| **WSC: Patchwork county (1)** | **10.95 (1.15-104.33)*** | **9.04 (1.49 – 54.81)*** |
| **WSC: University hospital (1)** | 0.60 (0.05 – 7.59) | 1.96 (0.24 – 16.30) |
| **WSC: HUS-area (1)** | 1.05 (0.07 – 15.69) | 0.50 (0.05 – 5.42) |
| **WSC: Population density** | 0.05 (0.00 – 17.53) | 0.93 (0.29 – 2.97) |
| **WSC: Number of municipalities** | 0.48 (0.11 – 2.04) | 0.35 (0.10 – 1.16) |
| **WSC: Budget surplus or deficit per person** | 1.36 (0.59 – 3.13) | 1.29 (0.64 – 2.60) |
| **Councilmembers from the municipality (%)** | 0.53 (0.19 – 1.44) | 0.39 (0.15 – 1.06) |
| **Largest party seat share (%)** | 0.91 (0.44 – 1.88) | 0.94 (0.48 – 1.70) |
| Variance of the random intercept: τ_00_ | 1.09 | 0.79 |
| ICC | 0.25 | 0.19 |
| AIC | 255.1 | 290.1 |
| *** = p < 0.001, ** = p < 0.01, * = p < 0.05  OR = Odds ratio, WSC = Wellbeing services county,  ICC = Intraclass correlation coefficient, AIC = Akaike information criterion | | |

**Annex 3. Full multilevel linear model of confirmed (1) and including unconfirmed (2) closures in a municipality as a continuous outcome variable.**

| **Full model comparison:** | **1. Number of confirmed closures** | **2. Number of all closures (including unconfirmed)** |
| --- | --- | --- |
|  | **Stand B (95 % CI)** | |
| **Population** | **-0.46 (-0.72 – -0.21)***** | -0.01 (-0.29 – 0.28) |
| **Population density (per km2)** | **0.26 (0.06 – 0.46)*** | **-0.30 (-0.53 – -0.08)**** |
| **Proportion of population aged 65 or older** | -0.06 (-0.23 – 0.11) | 0.02 (-0.17 – 0.21) |
| **Population growth (2013-2023)** | -0.13 (-0.31 – 0.04) | 0.05 (-0.15 – 0.24) |
| **Number of public health centers** | **1.06 (0.90 – 1.22)***** | **0.36 (0.18 – 0.54)***** |
| **Number of private clinics** | **-0.26 (-0.46 - -0.06)*** | 0.04 (-0.19 – 0.26) |
| **Medical desert index** | **0.12 (0.02 – 0.22)*** | -0.03 (-0.14 – 0.08) |
| **Previous closures in municipality (10 years) (1)** | -0.12 (-0.40 – 0.16) | 0.09 (-0.23 – 0.40) |
| **WSC: Patchwork county (1)** | 0.45 (-0.04 – 0.94) | 0.08 (-0.47 – 0.62) |
| **WSC: University hospital (1)** | -0.30 (-0.91 – 0.32) | 0.57 (-0.11 – 1.25) |
| **WSC: HUS-area (1)** | -0.09 (-0.73 – 0.56) | -0.16 (-0.88 – 0.55) |
| **WSC: Population density** | -0.22 (-0.45 – 0.01) | **0.59 (0.33 – 0.85)***** |
| **WSC: Number of municipalities** | -0.10 (-0.45 – 0.25) | -0.24 (-0.63 – 0.15) |
| **WSC: Budget surplus or deficit per person** | 0.08 (-0.12 – 0.27) | 0.00 (-0.21 – 0.22) |
| **Councilmembers from the municipality (%)** | 0.08 (-0.09 – 0.25) | -0.12 (-0.31 – 0.08) |
| **Largest party seat share (%)** | -0.00 (-0.17 – 0.16) | -0.01 (-0.20 – 0.17) |
| Variance of the random intercept: τ_00_ | 0.07 | 0.09 |
| ICC | 0.13 | 0.12 |
| *** = p < 0.001, ** = p < 0.01, * = p < 0.05  Stand B = Standardized beta coefficient, WSC = Wellbeing services county,  ICC = Intraclass correlation coefficient | | |
